# Association Between SARS-CoV-2 Mutations and Disease Severity Reveals Risk and Protective Effects Among Community-Sampled Patients in Israel

**DOI:** 10.64898/2026.01.26.26344903

**Authors:** Hagar Eliyahu, Noam Barda, Michal Mandelboim, Yaniv Lustig, Neta S. Zuckerman

## Abstract

SARS-CoV-2 mutations play a key role in viral evolution, in immune escape, and potentially in disease severity. However, the clinical impact of most mutations remains poorly understood, particularly across different variants. A historical observational study was conducted using SARS-CoV-2 whole-genome sequencing data linked to clinical metadata from 175,503 COVID-19 cases in Israel. The dataset was stratified into four variant-specific periods: B.1.1.7, B.1.617.2, BA.1, and BA.2. Logistic regression models were applied separately within each period to assess the association between individual mutations and the need for hospitalization, adjusting for age, gender, and time since vaccination. False discovery rate correction was used to account for multiple testing. A total of 18 SARS-CoV-2 mutations were significantly associated with COVID-19 severity, of which eight remained statistically significant after false discovery rate correction. Among these, two were associated with increased risk and six with reduced risk. Most were non-synonymous mutations located in functionally relevant regions such as the spike protein and non-structural proteins. This study provides a variant-stratified assessment of SARS-CoV-2 mutations associated with clinical severity, revealing both known and novel associations. The findings highlight the importance of integrating genomic and clinical data in public health surveillance and may inform future outbreak preparedness by identifying mutations with potential clinical impact.

## Introduction

Severe acute respiratory syndrome coronavirus 2 (SARS-CoV-2), responsible for the Coronavirus disease 2019 (COVID-19) pandemic, is a single-stranded RNA virus with ∼30,000 nucleotides which emerged in Wuhan, China, in 2019 [1]. The virus primarily spreads through respiratory droplets and can cause a spectrum of clinical manifestations ranging from mild symptoms to severe respiratory illness. As of today, more than 770 million cases have been confirmed worldwide, including over 4 million in Israel, with global deaths exceeding 6 million, 12,635 of which in Israel [2,3]. The severity of illness among hospitalized individuals varies widely, highlighting the need for tailored clinical management to optimize patient outcomes [1].

Since its emergence, SARS-CoV-2 has undergone various genetic mutations giving rise to distinct variants. These variants, characterized by specific mutations in the virus’ genomic sequences, have raised concerns due to their potential impact on transmissibility, severity of illness, and vaccine effectiveness. Some of these variants have been designated as variants of concern (VOC) by the World Health Organization due to specific properties they possess and their significant spread. For example, the alpha VOC, dominant worldwide in early 2021, and the delta VOC, dominant from mid-2021, were linked to higher transmissibility, increased disease severity and the ability to evade the immune response [5-8].

Previous studies have identified associations between specific SARS-CoV-2 mutations and clinical outcomes, including disease severity and mortality. Mutations in both structural and non-structural proteins have been implicated in either reducing or increasing the host response to infection. For example, changes in the nucleocapsid, spike glycoprotein, and various ORFs have been linked to more severe cases of COVID-19, while other mutations have been associated with milder presentations. Furthermore, variant-specific differences in mortality rates have been observed across different waves of the pandemic, highlighting the importance of genomic surveillance in understanding clinical trends [9–11].

Each SARS-CoV-2 VOC is defined by a distinct set of well-characterized mutations, many of which have been studied in relation to transmissibility and clinical outcomes. However, a wide range of additional, less characterized mutations may also arise within each SARS-CoV-2 VOC during infection and transmission. These under characterized mutations may influence disease severity or contribute to the emergence of new sub-lineages and transmission chains. In this study, we focused on these under characterized mutations outside the defining set for each variant and their potential associations with disease outcome.

## Materials and Methods

### Study Design

A historical observational analysis was conducted using SARS-CoV-2 whole-genome sequencing data linked to clinical metadata. The data were generated by the Israel Ministry of Health National Consortium for SARS-CoV-2 Sequencing (available in GISAID). Samples with sequencing coverage below 50 percent or incomplete clinical metadata, such as missing gender or implausible age values, were excluded. Following this filtering, the dataset included only unique patients.

The dataset was segmented into four VOC-specific subsets: B.1.1.7 VOC and sub-lineages - December 1, 2020 to March 30, 2021; B.1.617.2 VOC and sub-lineages - July 1, 2021 to October 30, 2021; BA.1 and sub-lineages - December 1, 2021 to March 30, 2022 ; BA.2 and sub-lineages - March 1, 2022 to May 30, 2022. The segmentation aligned with major COVID-19 epidemic waves in Israel during the study period (Figure 1). Clinical severity (hospitalized / non-hospitalized) was recorded for all cases.

**Figure 1.**
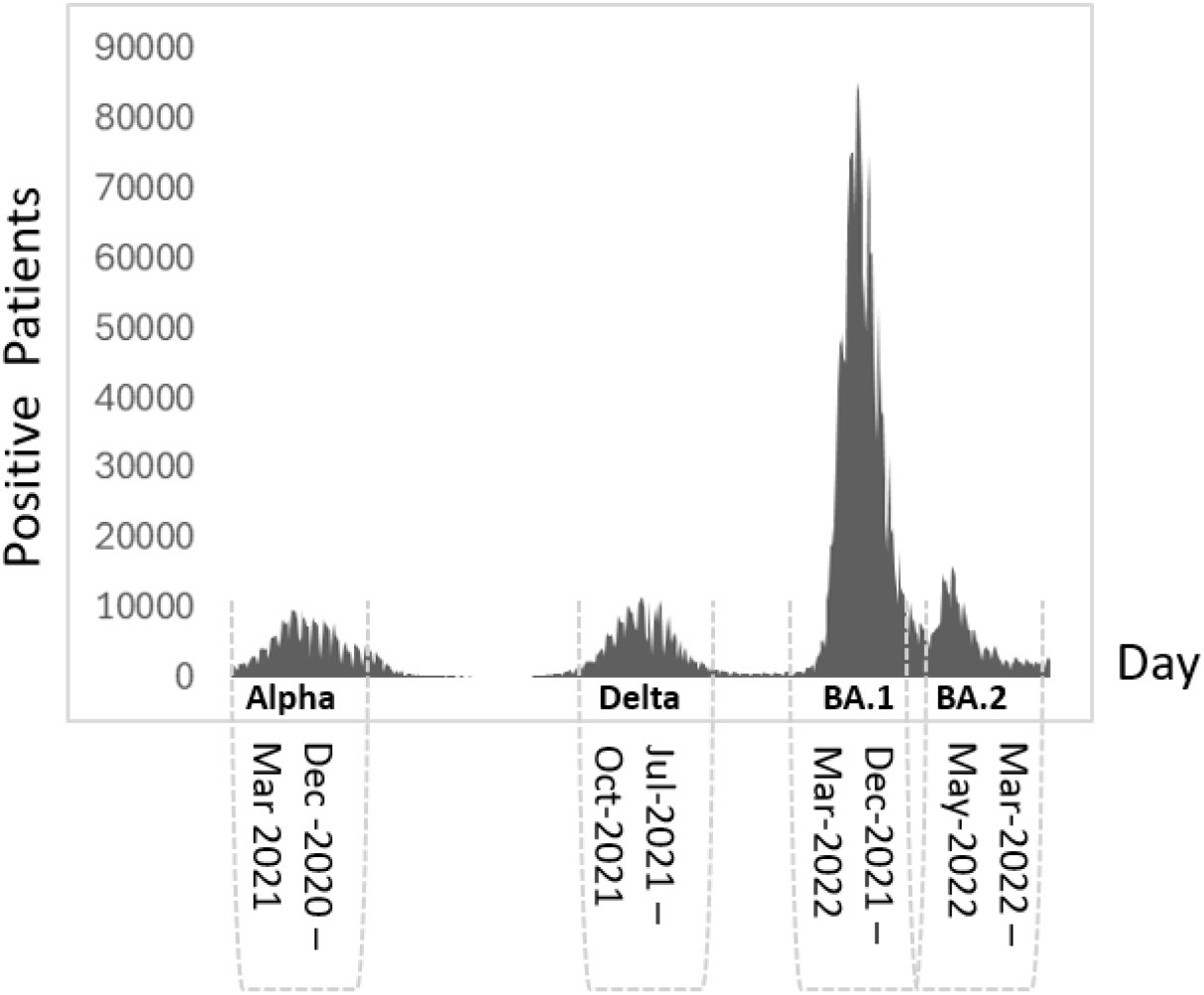
Dataset Segmentation by Variant Period and Hospitalization Status Epidemic curve showing the number of confirmed SARS-CoV-2 cases in Israel over time [2]. The periods corresponding to B.1.1.7 (December 2020–March 2021), B.1.617.2 (July– October 2021), BA.1 (December 2021–March 2022), and BA.2 (March–May 2022) are annotated on the timeline indicating the segmentation of the dataset by variant period.

### Data Collection and Processing

Clinical data including age, gender, days since last vaccination, and disease severity, were obtained from national health registries and linked to SARS-CoV-2 genomic data. The outcome of interest was disease severity, assessed based on whether an infection required hospitalization.

Whole-genome sequencing of SARS-CoV-2 samples was performed, and mutations were identified, using a custom bioinformatics pipeline, implemented in Bash and Python and executed on UNIX-based servers. Sequencing reads were aligned to the SARS-CoV-2 reference genome (NC_045512.2) using BWA-MEM (version: 0.7.17-r1188). Variant calling and consensus sequence generation were performed using iVar (version: 1.3.1), with a minimum depth threshold of five reads. Quality control metrics, including coverage statistics and read depth, were assessed using Samtools (version 1.11). Consensus sequences underwent multiple alignment with the reference genome using the MAFFT-based alignment module implemented in Augur (version: 15.0.2). Lineage classification was conducted with Nextclade (version 2.14.0) to assign SARS-CoV-2 variants. Mutations for each clinical sample relative to the reference sequence were identified and used for downstream statistical analysis. Both synonymous and non-synonymous mutations were included. mutations observed in at least 2% of cases were retained. CovSpectrum [15] was used to identify which mutations were prevalent in specific countries and variants and to assess their frequency within the population, based on global sequence data.

### Statistical Analyses

Logistic regression analyses were conducted separately for each mutation within each segment. Each model included disease severity as the dependent variable and was adjusted for age, gender, and days since last vaccination. P-values were corrected for multiple comparisons with the false discovery rate (FDR) method. Adjusted p-values under 0.05 were considered statistically significant. Odds ratios and 95% confidence intervals were derived from the model estimates. Analysis was performed using R, version 4.4.1.

### Phylogenetic Analysis

For the phylogenetic analysis, the Nextstrain Augur pipeline⍰[12] was employed. Time-resolved phylogenetic trees were constructed using IQ-TREE and TreeTime⍰[13,⍰14], employing the GTR substitution model. Final visualization was performed with Auspice. For each variant period, representative samples were selected, with all hospitalized cases included. Sampling was guided by mutations identified as statistically significant in the regression models. For each mutation, groups of individuals with and without the mutation were compared, and cases from both groups were included. After all relevant mutation patterns were included, additional cases were randomly sampled to reach a total of 1,000 sequences per variant period.

Co-occurring mutation sets, identified from the phylogeny, were evaluated using the same adjusted logistic regression model applied to single mutations, comparing individuals carrying all mutations in the set with those carrying none.

## Results

### Study Population and Dataset Segmentation to VOC-specific Subsets

A total of 205,300 confirmed COVID-19 cases were identified, of which 175,503 met the inclusion criteria. The dataset was segmented into four VOC-specific subsets: B.1.1.7 (n = 2,057); B.1.617.2 (n = 2,529); BA.1 (n = 12,926); BA.2 (n = 21,392).

### Association between mutations and disease severity

The distribution of mutations along the SARS-CoV-2 genome was similar across all variants and time periods, with the highest densities observed in the spike gene, followed by non-structural proteins (NSPs) and ORFs (Figure S1).

A total of 18 mutations significantly associated with COVID-19 severity were identified, with 8 remaining statistically significant after the FDR correction (Table 2A). Among the 8 mutations, 2 were associated with increased risk and 6 with reduced risk. Most mutations (6/8, 75%) were non-synonymous i.eleading to a change in the amino acid property (Table 2B). The hospitalization status distribution varied across the significant mutations (Figure S2).

All mutations were prevalent in Israel, while several were also prevalent in countries such as the USA, UK, Germany, Denmark, and Australia. The proportion of individuals carrying these mutations varied across variant periods. Mutations from the B.1.1.7 period were highly prevalent (51–58%), whereas BA.2 mutations showed wide variability, ranging from 2% to 77% (Table 2B).

### Associated mutations per variant

Within the B.1.17 variant, two mutations were associated with decreased disease severity. Spike:L5F (C21575T), a non-synonymous mutation altering a hydrophobic aliphatic residue to an aromatic one, found in 58.3% of the population; ORF7a:Q62* (C27577T), a nonsense mutation leading to an amino acid property change, found in 53% of the population. both primarily observed in Israel and the United States (Table 2).

No statistically significant mutations were identified within the B.1.617.2 or BA.1 variants.

Within the omicron BA.2 variant, six mutations were identified. Among the mutations associated with increased disease severity were NSP7:D77N (G12071A), a non-synonymous mutation altering the amino acid property group, found exclusively in Israel at a frequency of 5.6%, and Spike:N440K (T22882G), present in 35.0%, of the population. In contrast, mutations associated with reduced severity included Spike:S704L (C23677T), found in 3.9% of the population and resulting in an amino acid property shift, and NSP4:L264F (C9344T), the most prevalent mutation at 77.1%, also a non-synonymous mutation affecting the amino acid property group (Table 2).

### Transmission Landscape of SARS-CoV-2 Mutations Associated with Disease Severity in Israel

To represent the distribution of mutations identified as associated with disease severity among patients in Israel within the local transmission landscape, a phylogenetic analysis was conducted for each variant during its respective period of circulation. To capture the mutation landscape present at that time, a set of representative sequenced samples was randomly selected for phylogenetic analysis.

Within the B.1.17 variant, the mutations were present in most of the sequences. Sequences from patients with different hospitalization status were distributed uniformly across the phylogenetic tree, with no apparent clustering by clinical outcome (Figure 2). Given the co-occurrence of the two mutations in most sequences, a potential combined effect was evaluated. A logistic regression model was applied comparing cases carrying the mutations to those carrying none. The resulting odds ratio was similar to those obtained for each mutation individually (Table 2A).

**Figure 2.**
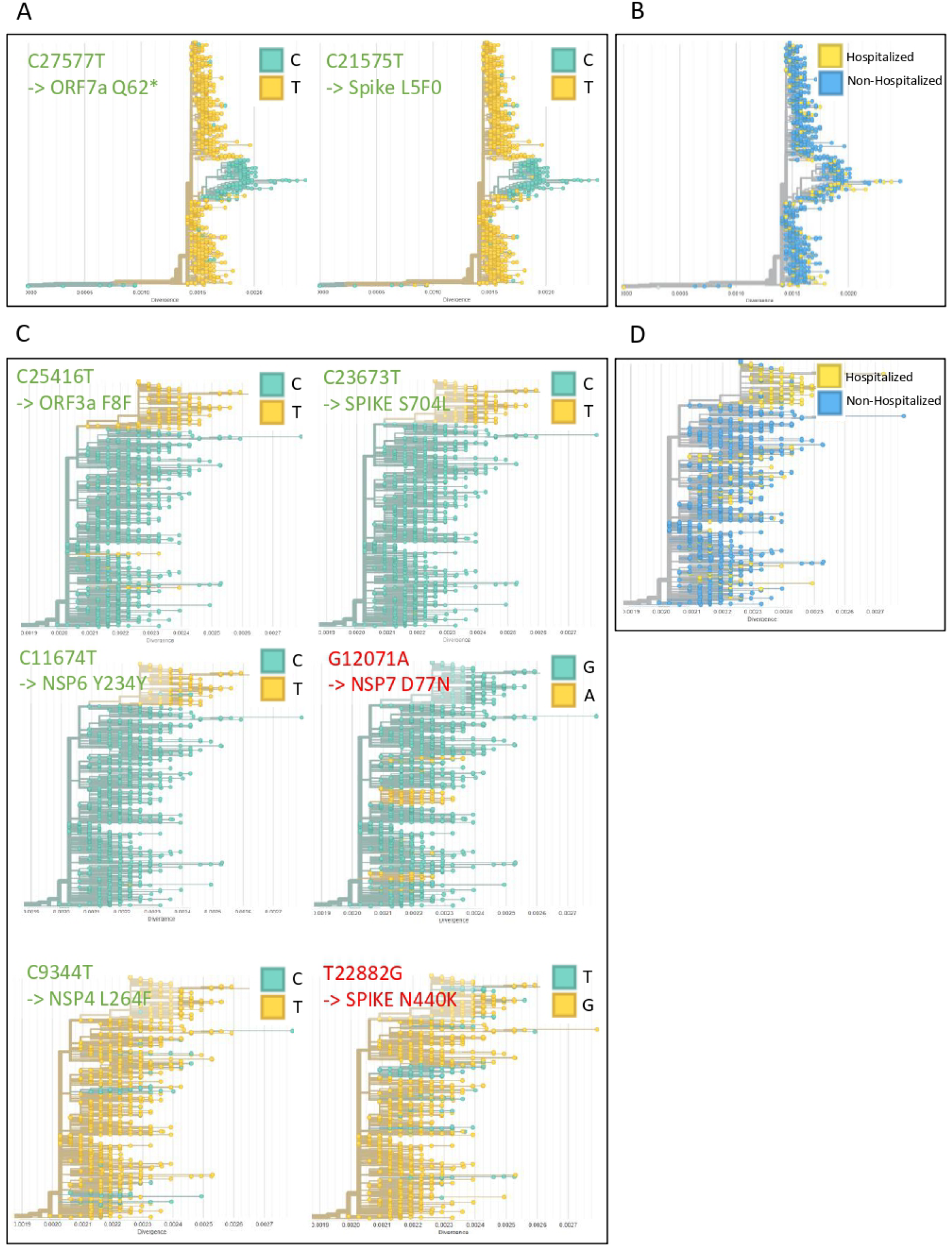
Phylogenetic trees of B.1.1.7 (A–B) and BA.2 (C–D) variants annotated by mutation presence and hospitalization status. (A, C) Trees annotated by the presence or absence of mutations associated with disease severity. (B, D) Trees annotated by hospitalization status (yellow: hospitalized; blue: non-hospitalized)

Within the BA.2 variant, three mutations associated with a protective effect (Spike:S704L, ORF3a:F8F, NSP6:Y234Y) were found to co-occur within a distinct sub-lineage, which contained very few hospitalized cases (Figure 2). A logistic regression model was applied to evaluate the combined effect of the three mutations, and the resulting odds ratio was similar to those obtained when each mutation was analyzed individually (Table 2A). Apart from this sub-lineage, sequences from hospitalized and non-hospitalized patients were interspersed throughout the phylogenetic tree across all variants (Figure 2).

The remaining four mutations within the BA.2 variant displayed varying phylogenetic patterns. NSP7:D77N was observed in three small clusters, Spike:N440K and NSP4:L264F appeared in multiple locations across the tree, with most sequences forming clusters, except for a few scattered cases. All of these mutations showed a similar distribution between hospitalized and non-hospitalized individuals.

## Discussion

This study assessed the association between under characterized SARS-CoV-2 variant mutations and COVID-19 disease severity in patients from Israel. A set of mutations associated with severity was identified across distinct SARS-CoV-2 variant periods. The predominance of non-synonymous mutations, particularly those altering amino acid properties, indicates potential functional impact on viral proteins. These patterns may reflect selective pressures acting on regions involved in host interaction, immune evasion, or replication efficiency.

Among the mutations identified within the B.1.17 variant, protective substitutions were observed in key viral regions. The Spike:L5F mutation, located in the signal peptide, may influence protein processing or trafficking and has been recurrently observed in local clusters, suggesting a possible selective advantage [13]. ORF7a:Q62*, which introduces a premature stop codon, may potentially impair immune evasion by truncating a protein involved in interferon suppression. Similar truncations have been linked to reduced replication and attenuated immune modulation [17]. The mutations were primarily observed in Israel, potentially reflecting the circulation of local sub-lineages during this early variant wave.

In both B.1.617.2 and BA.1, no mutations remained statistically significant after FDR correction. This likely reflects the distribution of clinical outcomes within our study population, rather than a true absence of relevant viral changes. The proportion of hospitalized cases was markedly lower in BA.1 (1.5%) and B.1.617.2 (2.3%) compared with B.1.1.7 (8.7%) and BA.2 (3.5%) (Table 1). As our models evaluate associations with disease severity, the number of severe outcomes directly affects statistical power. Although B.1.617.2 and BA.1 contributed large numbers of total cases, their low frequency of hospitalizations limited the ability to detect statistically significant associations. In contrast, B.1.1.7 and BA.2, where severe cases were more frequent, yielded multiple significant associations.

**Table 1.**
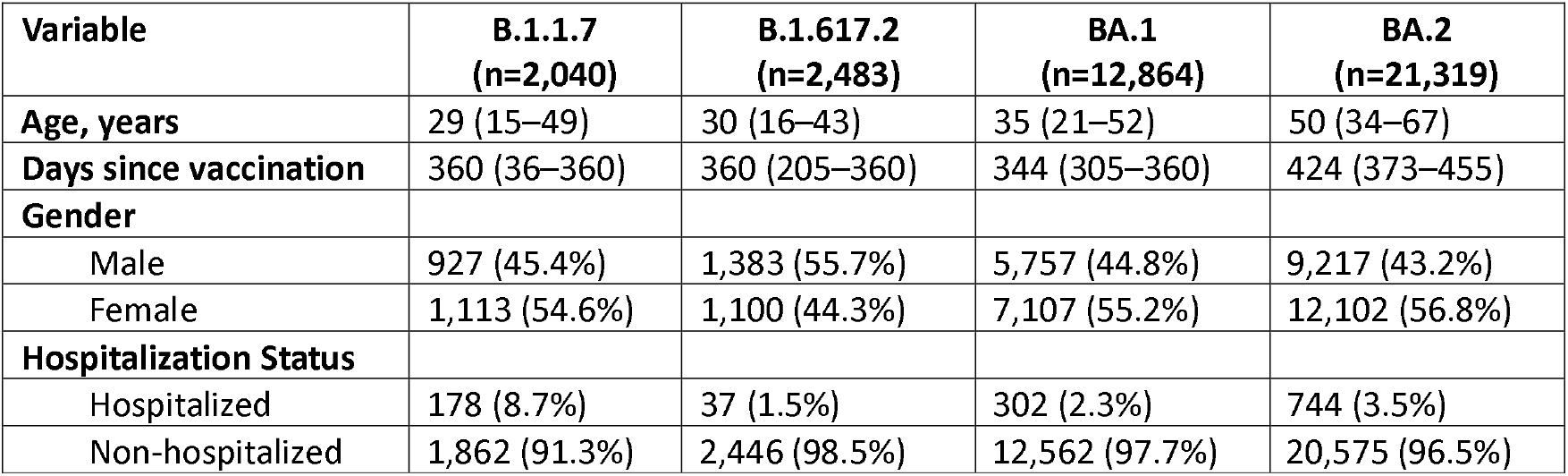
Demographic and clinical characteristics of study participants by SARS-CoV-2 variant. Continuous variables are presented as median (interquartile range, Q1–Q3); categorical variables are presented as number (percentage). Days since vaccination refers to the number of days between the last vaccine dose and the date of positive SARS-CoV-2 test.

| Variable | B.1.1.7<br>(n=2,040) | B.1.617.2<br>(n=2,483) | BA.1<br>(n=12,864) | BA.2<br>(n=21,319) |
| --- | --- | --- | --- | --- |
| Age, years | 29 (15–49) | 30 (16–43) | 35 (21–52) | 50 (34–67) |
| Days since vaccination | 360 (36–360) | 360 (205–360) | 344 (305–360) | 424 (373–455) |
| Gender |  |  |  |  |
| Male | 927 (45.4%) | 1,383 (55.7%) | 5,757 (44.8%) | 9,217 (43.2%) |
| Female | 1,113 (54.6%) | 1,100 (44.3%) | 7,107 (55.2%) | 12,102 (56.8%) |
| Hospitalization Status |  |  |  |  |
| Hospitalized | 178 (8.7%) | 37 (1.5%) | 302 (2.3%) | 744 (3.5%) |
| Non-hospitalized | 1,862 (91.3%) | 2,446 (98.5%) | 12,562 (97.7%) | 20,575 (96.5%) |

**Table 2.**
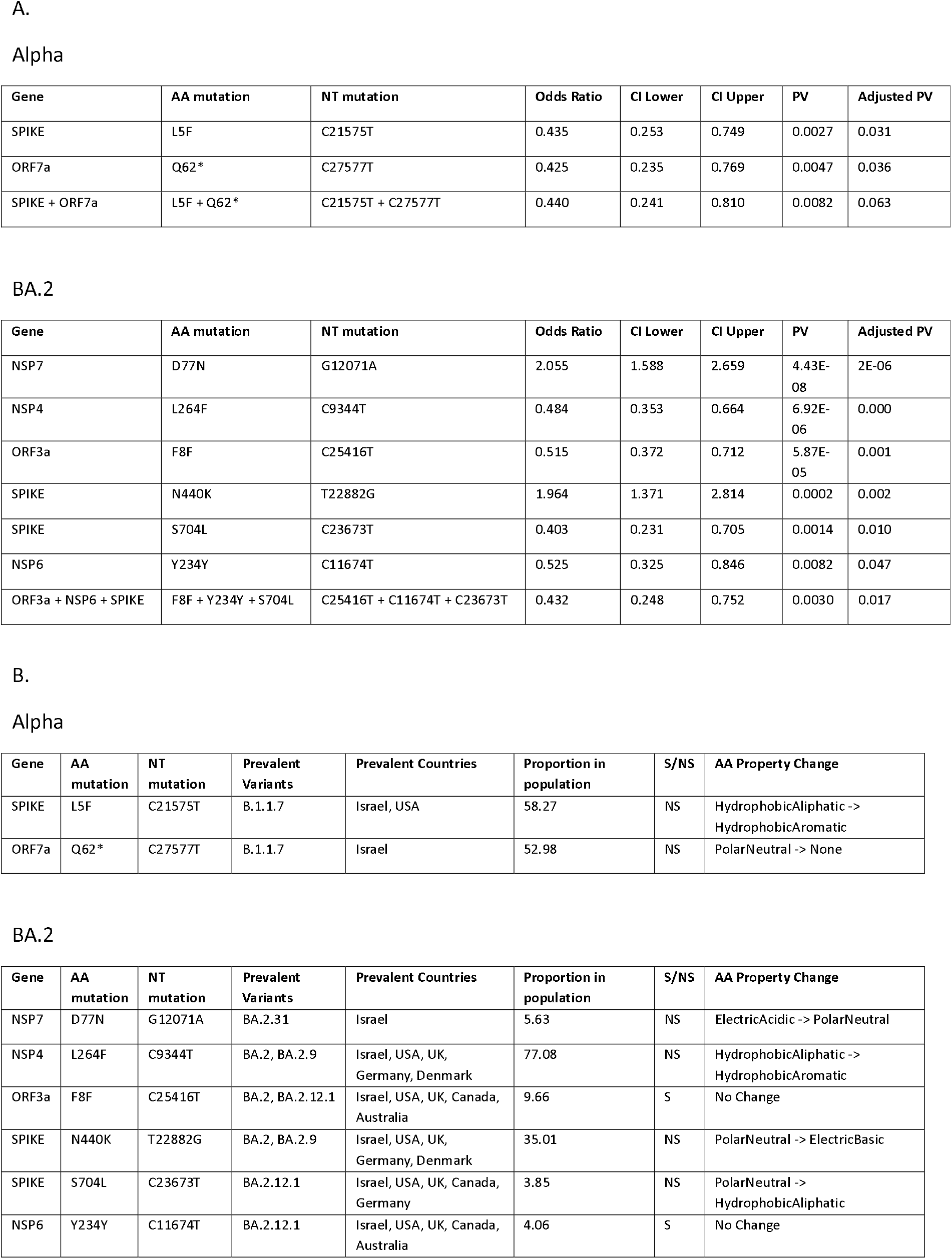
Summary of Mutations Associated with COVID-19 Severity. A. Logistic regression model results. For each mutation, the gene, amino acid (AA) change, and corresponding nucleotide (NT) change are shown alongside logistic regression estimates. Models were adjusted for age, gender, and days since vaccination. Odds ratios (OR), 95% confidence intervals (CI), and both raw and FDR-adjusted p-values are reported. B. Characteristics of significant and notable mutations. For each mutation, the dominant countries and estimated global proportion in the population were obtained from the CovSpectrum platform [15]. Mutation type - S (synonymous) or NS (non-synonymous), and amino acid property change (if applicable) are also listed.

| Gene | AA mutation | NT mutation | Odds Ratio | CI Lower | CI Upper | PV | Adjusted PV |
| --- | --- | --- | --- | --- | --- | --- | --- |
| SPIKE | L5F | C21575T | 0.435 | 0.253 | 0.749 | 0.0027 | 0.031 |
| ORF7a | Q62* | C27577T | 0.425 | 0.235 | 0.769 | 0.0047 | 0.036 |
| SPIKE + ORF7a | L5F + Q62* | C21575T + C27577T | 0.440 | 0.241 | 0.810 | 0.0082 | 0.063 |

BA.2
| Gene | AA mutation | NT mutation | Odds Ratio | CI Lower | CI Upper | PV | Adjusted PV |
| --- | --- | --- | --- | --- | --- | --- | --- |
| NSP7 | D77N | G12071A | 2.055 | 1.588 | 2.659 | 4.43E-08 | 2E-06 |
| NSP4 | L264F | C9344T | 0.484 | 0.353 | 0.664 | 6.92E-06 | 0.000 |
| ORF3a | F8F | C25416T | 0.515 | 0.372 | 0.712 | 5.87E-05 | 0.001 |
| SPIKE | N440K | T22882G | 1.964 | 1.371 | 2.814 | 0.0002 | 0.002 |
| SPIKE | S704L | C23673T | 0.403 | 0.231 | 0.705 | 0.0014 | 0.010 |
| NSP6 | Y234Y | C11674T | 0.525 | 0.325 | 0.846 | 0.0082 | 0.047 |
| ORF3a + NSP6 + SPIKE | F8F + Y234Y + S704L | C25416T + C11674T + C23673T | 0.432 | 0.248 | 0.752 | 0.0030 | 0.017 |

B. Alpha
| Gene | AA mutation | NT mutation | Prevalent Variants | Prevalent Countries | Proportion in population | S/NS | AA Property Change |
| --- | --- | --- | --- | --- | --- | --- | --- |
| SPIKE | L5F | C21575T | B.1.1.7 | Israel, USA | 58.27 | NS | HydrophobicAliphatic -> HydrophobicAromatic |
| ORF7a | Q62* | C27577T | B.1.1.7 | Israel | 52.98 | NS | PolarNeutral -> None |

Table 2. Summary of Mutations Associated with COVID-19 Severity
| Gene | AA mutation | NT mutation | Prevalent Variants | Prevalent Countries | Proportion in population | S/NS | AA Property Change |
| --- | --- | --- | --- | --- | --- | --- | --- |
| NSP7 | D77N | G12071A | BA.2.31 | Israel | 5.63 | NS | ElectricAcidic -> PolarNeutral |
| NSP4 | L264F | C9344T | BA.2, BA.2.9 | Israel, USA, UK, Germany, Denmark | 77.08 | NS | HydrophobicAliphatic -> HydrophobicAromatic |
| ORF3a | F8F | C25416T | BA.2, BA.2.12.1 | Israel, USA, UK, Canada, Australia | 9.66 | S | No Change |
| SPIKE | N440K | T22882G | BA.2, BA.2.9 | Israel, USA, UK, Germany, Denmark | 35.01 | NS | PolarNeutral -> ElectricBasic |
| SPIKE | S704L | C23673T | BA.2.12.1 | Israel, USA, UK, Canada, Germany | 3.85 | NS | PolarNeutral -> HydrophobicAliphatic |
| NSP6 | Y234Y | C11674T | BA.2.12.1 | Israel, USA, UK, Canada, Australia | 4.06 | S | No Change |

Several mutations observed within the BA.2 variant were associated with decreased disease severity and mapped to functionally relevant regions of the SARS-CoV-2 genome. One such mutation, Spike:S704L, is characteristic of the globally circulating BA.2.12.1 sub-lineage [21]. It is located near the S1/S2 cleavage site, a region critical for spike protein processing and viral entry [22]. Although BA.2.12.1 demonstrated enhanced global transmissibility [24], the observed protective association of S704L suggests that increased spread does not necessarily translate to greater clinical severity. Another protective mutation, NSP4:L264F, lies within a large luminal loop of NSP4, a protein essential for the formation of double-membrane vesicles that support viral replication [26]. This mutation has been detected across multiple Omicron subvariants, excluding BA.1. Its recurrence in these lineages may indicate a functional advantage that promotes efficient viral replication while maintaining a relatively mild clinical presentation, reflecting a potential balance between viral fitness and reduced disease severity.

By contrast, the NSP7 D77N mutation, a common substitution in the nonstructural protein 7 gene [25], was associated with increased disease severity. NSP7 is a critical cofactor of the RNA-dependent RNA polymerase (RdRp) complex, and destabilization of this interaction may impair replication fidelity. The SARS-CoV-2 RdRp complex (nsp12/7/8) has already been shown to exhibit unusually low base substitution fidelity in the absence of proofreading [26], and further reductions in fidelity could increase viral diversity, facilitate immune evasion, and ultimately contribute to more severe disease outcomes. Another mutation associated with increased risk, Spike N440K, is located within the receptor-binding domain of the spike protein and has previously been linked to immune escape and reduced susceptibility to neutralization by monoclonal antibodies [20].

Two synonymous mutations were observed within the BA.2 variant group: ORF3a F8F and NSP6 Y236Y. Although these substitutions do not alter the encoded amino acids and their functional impact has not been reported in the literature, the protective association observed for these mutations may suggest they serve as markers of broader genomic contexts, potentially reflecting linkage to functional mutations or lineage-defining signatures.

Across the set of significant mutations, the distribution of hospitalization status varied (Figure S2). This finding indicates that associations with disease severity were not restricted to mutations highly enriched in either hospitalized or non-hospitalized individuals, supporting the relevance of investigating each mutation individually.

Phylogenetic analysis of the SARS-CoV-2 transmission landscape in Israel revealed that most cases had a similar distribution of hospitalization status across the phylogenetic tree, regardless of whether the defining mutations were associated with increased or decreased risk. This suggests that disease severity is not solely determined by infection with a specific mutation or sub-lineage. These findings highlight the importance of applying statistical models that adjust for potential confounders. In our analysis, age, vaccination status, and gender were included as covariates, allowing the associations observed between specific mutations and hospitalization to be interpreted as independent of these factors.

Notably, within the B.1.1.7 and BA.2 variant periods, clusters of co-occurring mutations were identified that had a similar protective effect. The interaction analyses performed to compare the combined vs. individual effect of these mutations showed a similar protective association in both, suggesting that these mutations may define circulating strains associated with reduced disease severity.

This study has several limitations. First, the relatively low number of hospitalized patients within the B.1.617.2 and BA.1 variant groups reduced statistical power, thereby preventing the detection of mutations reaching statistical significance in these groups. Second, although models were adjusted for key covariates such as age, vaccination status, and gender, unmeasured or unknown confounders may have influenced the observed associations. Third, hospitalization status may be subject to misclassification in clinical records, as some individuals may have been admitted for reasons unrelated to COVID-19 but were nonetheless recorded as COVID-19 hospitalizations due to incidental positive testing [28].

This study highlights the importance of monitoring specific SARS-CoV-2 mutations rather than focusing solely on viral lineages. While some mutations were associated with increased risk of severe disease and others appeared protective, their clinical impact varied depending on the variant in which they appeared. These findings highlight the need to evaluate mutations in their specific genomic and epidemiological context, as the same substitution may lead to different outcomes across strains. Additionally, by adjusting for confounders such as age, gender, and vaccination status, this analysis revealed associations that may not be apparent through phylogenetic clustering alone. As the virus continues to evolve, mutation-level surveillance, paired with careful clinical interpretation, will be key to anticipating disease outcomes and informing public health decisions.

## Data Availability

All data produced are available online at Gisaid

## Author Contributions

Conceptualization, H.E., N.Z and Y.L.; Methodology, H.E., N.B., N.Z and Y.L..; Validation, H.E., N.B., N.Z and Y.L.; Formal Analysis, H.E..; Investigation, H.E..; Resources, M.M.; Writing – Original Draft Preparation, H.E. and N.Z.; Writing – Review & Editing, H.E., N.Z and Y.L.; Visualization, H.E.; Supervision, N.Z and Y.L.; Project Administration, N.Z and Y.L.

## Institutional Review Board Statement

The study was conducted in accordance with the Declaration of Helsinki, and approved by the Institutional Review Board of Sheba Medical Center (approval number SMC-1194-24).

## Informed Consent Statement

Patient consent was waived because the study used residual clinical samples and anonymized clinical data.

## Acknowledgments

We would like to thank the Israel Ministry of Health Public Health Services and Health Intelligence, the Ministry of Health Center for Knowledge and Information, and the Israel National Consortium for SARS-CoV-2 Sequencing.

