## supplementaty for "Association Between SARS-CoV-2 Mutations and Disease Severity Reveals Risk and Protective Effects Among Community-Sampled Patients in Israel"

Figure S1. Distribution of SARS-CoV-2 Mutations Identified Across Genes

(A) Number of mutations per gene normalized to gene length (mutations per kilobase).

(B) Mutation count per gene.

**A** **B**

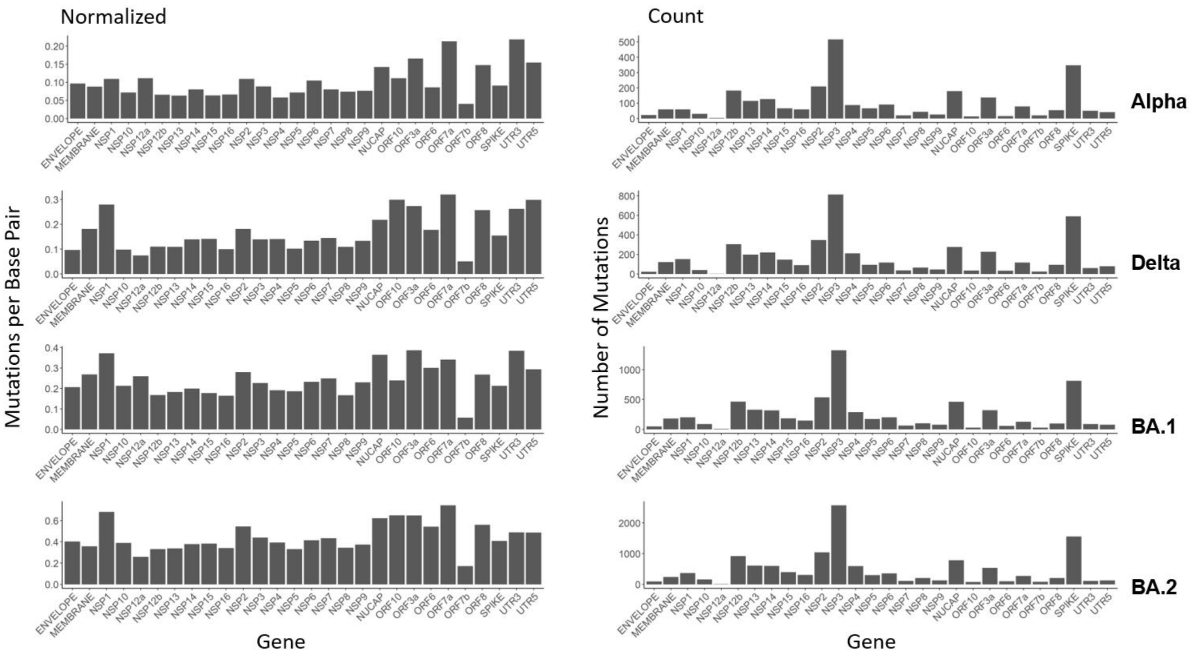

Figure S2. Hospitalization status by presence or absence of mutations associated with disease severity.

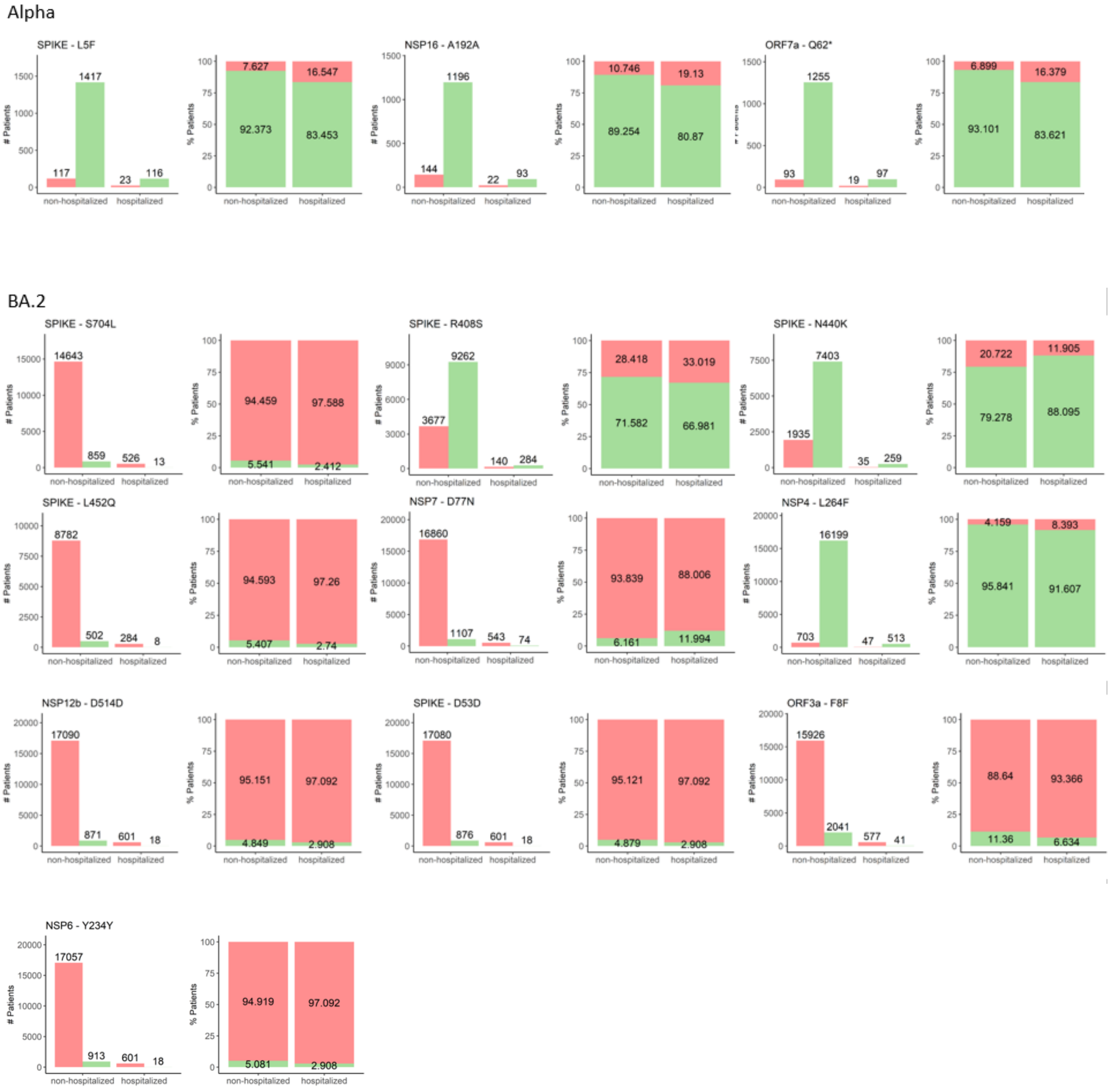
